# Mental Health in Ireland During the Covid Pandemic: Evidence from Two Longitudinal Surveys

**DOI:** 10.1101/2022.12.12.22283343

**Authors:** David Madden

## Abstract

Using data from the Growing Up in Ireland Covid survey, this study examines the evolution of mental health as measured in December 2020, nine months into the pandemic, compared to observations pre pandemic for two cohorts of people. A deterioration in mental health was observed for both cohorts and particularly for younger women of the 1998 cohort. The increase in the rate of depression predominantly occurred due to an overall decline in mental health rather than being concentrated amongst those already vulnerable (in the sense of being near the depression threshold). There was little, if any, change in the socioeconomic gradient associated with mental health and virtually no gradient at all was observed pre or post pandemic for the 1998 cohort. Mobility analysis revealed that not only did females from the 1998 cohort show greater transitions into depression, they also appeared to transition into more extreme levels of depression.

**JEL Codes:** I14, I31.

This research did not receive any specific grant from funding agencies in the public, commercial, or not-for-profit sectors.

**Abstract:** *Background:* The Covid pandemic arrived in Ireland on February 29, 2020. In the following weeks various restrictions were introduced to stem the spread of the disease. Anxiety over the spread of the disease and over the restrictions introduced had an adverse effect upon mental health. This study examines the change in mental health for two groups: young adults aged around 23 at the time of onset of Covid (the 1998 cohort) and a sample of principal carers (PCs) of children who were aged 13 at the onset of Covid (the 2008 cohort).

*Methods:* Data were obtained from the two cohorts of the longitudinal Growing Up In Ireland (GUI) survey. The sample included 1953 young adults (from the 1998 cohort) and 3547 principal carers (from the 2008 cohort). Mental health as measured by the Centre for Epidemiological Studies Depression - 8 scale was obtained for the last pre-Covid wave and for the Covid wave (surveyed in December 2020). Observations for which CES-D8 was not available in either pre or post Covid waves were excluded. Post-Covid sampling weights were applied.

*Results:* Relative to the last pre-Covid survey, mental health, as measured by CES-D8 deteriorated for both the young adults of the 1998 cohort and the PCs of the 2008 cohort. For young adults, the deterioration was more pronounced for females. There was no observable socioeconomic gradient for poor mental health amongst young adults, both pre and post Covid. For mothers from the 2008 cohort, a gradient was observed pre-Covid with poorer mental health for lower-income, less well-educated mothers. This gradient was less pronounced post-Covid, the levelling-off arising from a greater deterioration in mental health for higher-income and better-educated PCs.

*Conclusion:* Both observed cohorts showed a significant deterioration in mental health post Covid. For young adults the effect was more pronounced among females.

## Introduction

The Covid 19 Pandemic officially arrived in Ireland on February 29, 2020 with the first confirmation of a positive case. Over subsequent weeks various restrictions were introduced to stem the spread of the disease (becoming collectively known as the “lockdown”). These included the closure of all educational establishments and childcare facilities, the banning of various sporting and cultural events and then on March 27, everyone, apart from providers of essential care and services, were advised to stay at home apart from essential visits (e.g. to the supermarket) and exercise within a 2km radius. There was a ban on non-essential travel and on meeting people outside the immediate household.

As Covid cases declined over the summer of 2020 there was a gradual removal of the most severe of these restrictions, but an upsurge in autumn 2020 led to a reimposition of high level restrictions in October. As the second wave of Covid receded there was an easing of restrictions from early December with the opening of non-essential shops and services, and by December 18 limited within-country travel and household visits were permitted. However, there was a significant resurgence of cases in the immediate run-up to and aftermath of Christmas and severe restrictions were again imposed in January 2021.

The various lockdowns were successful in limiting the spread of Covid but these measures were not without their own costs. The most severe restrictions inevitably led to a reduction in economic activity and the reduction in human contact and the hardship imposed by social distancing raises concerns about the possible adverse mental health effects. We investigate the latter phenomenon in this paper. The landmark *Growing Up in Ireland* (GUI) longitudinal study carried out online surveys, including questions on mental health, for both the infant (2008) and child (1998) cohort for most of the month of December (the 2008 cohort survey began on December 4 and the 1998 cohort survey began on December 11, both surveys ending at the end of December). The 1998 cohort was aged between 22 and 23 at the time of this survey. The 2008 cohort was aged 12 at the time of the survey and their PCs (in almost all cases the biological mothers) were also surveyed. This group ranged in age from 33 to 55. No data were collected for the PCs of the older cohort.

GUI thus provides a snapshot of mental health for these groups at a time when restrictions were still relatively tight and had been ongoing for nearly nine months. Critically, since this is a longitudinal study we also have similar information *for the same people* for a period *before* Covid (for the 2008 cohort between June 2017 and February 2018, and for the 1998 cohort between August 2018 and June 2019).

These groups are by no means representative of the whole nation, though the 2008 and 1998 cohorts are representative of those people born in those specific periods. Nevertheless, what may be lacking in national representativeness is compensated for by having the same measure, for the same observations before and during the pandemic. We will investigate how mental health changed over this period, whether this change differed by gender and socioeconomic status and also the pattern of mobility for those people who did experience changes in their mental health.

A number of studies have examined the impact of Covid restriction on various measures of mental health or well-being. Aknin et al (2022) found that the restrictions introduced at the initial stages of Covid (around March 2020) impacted upon subjective well-being and psychological distress, but that much of this impact had abated by June 2020. Banks and Xu (2020) found substantial population level effects in a survey carried out in April 2020, with particularly marked effects upon young adults and women. Davillas and Jones (2021) also found substantial effects early in the pandemic but that much of this had gone by July 2020. Hajek et al (2022a, 2022b) investigate anxiety and depression across seven European countries during the later stages of Covid (from November 2020 to September 2021) and find highest rates for young people, aged 18-29. Tofolutti et al (2022) looked at the impact of Covid policies on mental well-being for 28 European countries from April 2020 to March 2021. They found that restrictions on international travel, private gatherings and contact tracing were associated with reductions in mental well-being of up to 4 per cent, with greater effects for females and those living with younger children. However, they do not have information on mental well-being before Covid.

In terms of studies for Ireland, Hyland et al (2021) find no evidence of an increase in mental health problems for Irish adults during the first year of the pandemic. It is noticeable however that they have no measures of mental health for their sample before the pandemic, so while mental health may not have deteriorated subsequent to the arrival of the pandemic, their analysis cannot tell if the pandemic caused an immediate deterioration in mental health.

Smyth and Murray (2022) use the GUI Covid module to investigate mental health for the 2008 cohort. However, since they do not have a consistent measure of mental health pre and post Covid their study is unable to explicitly examine how mental health changed with the pandemic. They analyse the association between the level of mental health in December 2020 and various factors such as family financial and education resources and restrictions on social activities. They find that lower mental health for the 2008 cohort was associated with a fall in family income arising from the pandemic (as opposed to family income pre-pandemic) and also with lower educational resources in terms of access to a computer and/or a quiet place to study.

The advantage provided by our study is that of a larger dataset than has been the case for other studies for Ireland (excepting Smyth and Murray) and critically the availability of a measure of mental health for the same people both before and after Covid. This is in contrast to other studies which have only followed mental health after the onset of the pandemic. The richness of our dataset also allows us to investigate whether the socioeconomic gradient of mental health issues differed pre and post pandemic and we are also able to examine transitions into and out of depression.

## Methods

### Study Design

This study is a cross-sectional, secondary analysis of quantitative data that were collected as part of the child cohort of the Growing Up in Ireland (GUI) study. The GUI is a nationally-representative sample of children living in Ireland.

### Ethics

The same ethical protocols govern all waves of the GUI survey (Kelly et al., 2021). The relevant pieces of legislation are: (i) the Statistics Act, 1993 which provides a strong legal basis for the protection of all information collected in Growing Up in Ireland against unlawful disclosure; (ii) the Children First Act 2015 -which is designed to raise awareness of child abuse and neglect and to ensure an appropriate response to it; (iii) the Data Protection Acts 1988, 2003, 2018 which clarify the rights of persons with respect to personal data that is processed concerning them. The Irish Department of Children, Equality, Disability, Integration and Youth and the Irish Central Statistics Office are joint Data Controllers for the survey; (iv) the Data Protection Act 2018 (Section 36(2)) (Health Research) Regulations 2018 which provides for additional safeguards when processing data of a sensitive nature such as health data.

The following principles were also applied: (i) Informed consent, with the providing of information on the purpose of the study, the type of information gathered and what will be involved for participants (ii) Reporting concerns regarding risks to children with a protocol for reporting any incidents and for handling these appropriately (iii) Confidentiality of information provided, which is a legal requirement under the Statistics Act (iv) Avoidance of harm (including embarrassment/distress) (v) Instruments and protocols were reviewed by an independent Research Ethics Committee.

Written consent was obtained for all participants in the study. Assent was also sought from the children who participated. Re-analysis of the GUI dataset does not require additional ethical approval in accordance with the Central Statistics Office of Ireland who hold the GUI dataset.

### Study Population

The data consist of two cohorts of Irish children and their PCs, the 2008 Cohort born in the period December 2007-June 2008 and the 1998 Cohort born in the period November 1997-October 1998 (Thornton et al, 2013 and Williams et al, 2009). The specific GUI data analysed is the last available (pre-Covid) waves of the GUI 2008 and 1998 Cohorts (collected in 2017/18 and 2018/19 respectively) and the Covid survey which was sent out in December 2020.

The latest pre-Covid wave of the 2008 Cohort consisted of 8032 children and their PCs and the original sampling frame used was the Child Benefit Register. This is a universal payment (made on behalf of all children regardless of socioeconomic status) and payment is made directly to the principal carer of the child (most typically the resident mother or step mother) and must be claimed within six months of the child being born, in the six months after the child becomes a member of the family or six months after the family become resident in Ireland. In the case of the 1998 Cohort the latest pre-Covid wave consisted of 5190 young adults and the original sample frame was the national primary school system, with 910 randomly selected schools participating in the study.

The following exclusions were placed on the data: for both cohorts only a balanced panel was used i.e. the study children (or young adults) and their principal carers who responded to the Covid survey and the last pre-Covid survey. In addition, observations where the questions on mental health were not answered were also excluded. As the mental health data collected in the last pre-Covid survey and the Covid survey for the children of the 2008 Cohort was not the same, these children were also excluded from the analysis. Following these exclusions, this left two study groups: the young adults from the 1998 Cohort (1953 observations, 1246 females and 707 males), and the PCs from the 2008 Cohort (3547 observations, 3457 females and 90 males). Since attrition from the GUI dataset is not random (McCrory et al, 2013) sampling weights from the Covid survey were used in all the analysis.

### Measurements

The measure of mental health we use for both of these study groups is the CES-D8 scale (Melchior et al 1993). The original version of the CES-D scale had 20 items and has been used extensively across the world and has featured in many published journal articles. There are also shorter versions of the measure which take less time to administer but are still regarded as reliable measures of depression. One of these is the CES-D8 and this is the version which is measured in GUI.

The CES-D8 measure consists of eight statements regarding how the respondent was feeling in the past week (e.g. “I felt depressed”, “I felt fearful” etc). The respondent then indicates whether they experienced this feeling rarely/none of the time, some or a little of the time, occasionally or a moderate amount of the time or most or all of the time. Answers are coded 0, 1, 2 or 3 respectively, so that the minimum score possible is 0 and the maximum is 24. Higher scores indicate worse mental health and a score at or above 7 is regarded as indicating depression (Devins et al, 1988). Our data is truncated at 13 (i.e. all CES-D8 scores greater than or equal to 13 are coded as 13) and hence the bulk of our focus is on rates of depression rather than actual CES-D8 scores..

As well as analysing how CES-D8 changes pre and post Covid we also examine its relationship to socioeconomic status (SES). We use two measures of SES. For the 1998 cohort of young adults we use the highest level of education attained by their principal carer. We break down education into four categories: (1) up to and including completion of lower secondary schooling (2) completion of all secondary schooling (3) obtaining a post-secondary school diploma or cert and (4) completion of third level education. We also include a category where this data is missing. We choose not to use the highest level of education obtained by the young adults themselves as many of the 1998 cohort will not have completed education, or may have had their education disrupted by Covid. For the PCs of the 2008 cohort we use their highest level of education obtained, using the same categories as above.

The second measure of SES we use is equivalized after tax household income, as measured in the last pre-Covid wave of GUI as this data was not collected in the Covid survey. This is calculated via the answer to questions on total net household income from all members and sources after deductions for income tax and social insurance. If households cannot give an exact figure then they answer a sequence of questions where they are presented with cards where they select the range into which they believe that family income falls. The measure of SES gradient we use for equivalized income is rank based and it seems reasonable to assume that even if households exact level of income is not measured with complete accuracy that the *ranking* of households by income will be less prone to error.

### Statistical Analysis

The following statistical analysis was carried out:

The distribution of CES-D8 scores and the fraction indicated as depressed are provided for both study groups. This analysis is stratified by gender for the 1998 cohort and by the SES measure of education for both study groups.

The Shapley decomposition is applied to the change in the fraction depressed (Kolenikov and Shorrocks, 2005). Suppose we characterise our measure of depression as *D* = *D*(*μ, L, CESD*^*^) where *μ* is the average level of CES-D8, *L* is the Lorenz curve for the distribution of CES-D8 and *CESD\** is the critical depression threshold (note that the cumulative distribution function for CES-D8 will be completely characterised by its mean and Lorenz curve).

If subscripts “0” and “1” refer to the two time periods in question, then the change in depression over time *D*_1_ ― *D*_0_ can be written as

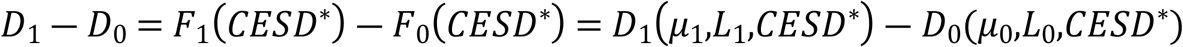

where

*F*_*i*_ is the cumulative distribution function for period “i”. This can then be decomposed into growth and redistribution effects denoted by *D*(*μ*_1_,*L*_0,_ *CESD*^*^) ―*D*(*μ*_0_,*L*_0_,*CESD*^*^) and *D* (*μ*_1_,*L*_1,_ *CESD*^*^) ―*D*(*μ*_1_,*L*_0_,*CESD*^*^) respectively.

However, as is the case with any path dependence type analysis, the choice of which configuration to use as the base period is arbitrary. In the above formulation we calculate the marginal effect of the change in mean CES-D8 with the distribution held constant at the *initial* configuration. However, we calculate the marginal impact of redistribution holding mean CES-D8 constant at the *final* configuration. We could just as easily have carried out a decomposition with the base periods changed and there is no logical reason for preferring one configuration over another. Following the approach outlined in Kolenikov and Shorrocks (2005) we take the average of the two effects respectively thus giving a growth effect of

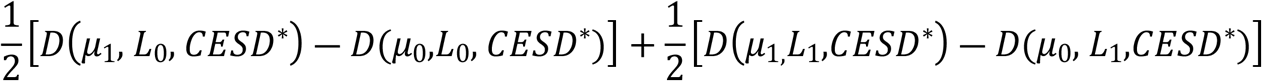

and a redistribution effect of

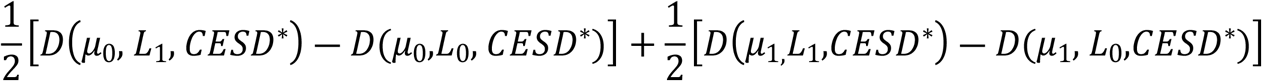

These two expressions are the growth and distribution components for a two-way Shapley decomposition of the change in the rate of depression. The Shapley decomposition arises from the classic co-operative game theory problem of dividing a pie fairly. The solution is that each player is assigned her marginal contribution averaged over all possible coalitions of agents. The interpretation here was to consider the various n factors which contribute together to determine the change in the value of an indicator such as depression and then assign to each factor the average marginal contributions taken over the n! possible ways in which the factors may be removed in sequence. Since we have two factors (n=2, growth and distribution) we have 2!=2 possible routes. The decomposition is always exact as the factors are treated symmetrically.

To investigate the gradient of mental health/depression with respect to SES, we examine how CES-D8 scores and depression rates vary by education of principal carer. To examine the gradient with respect to equivalised income we use the concentration index (Kakwani et al, 1997). Assume that the CES-D measure for individual i is given by *CESD*_*i*_. Then if *r*_*i*_ is the fractional rank of individual i in the income distribution (or whatever ranking variable is being used), the concentration index for CES-D is given by

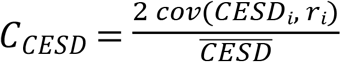 where 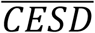 is the mean value of CES-D. ***C***_*CESD*_ can take on a value from -1 to +1, where a negative (positive) value indicates that CES-D takes on higher values among the relatively poor (rich). In the case of a binary measure (e.g. depressed/non-depressed) a normalisation must be applied to the index (since the bounds would not be -1 and +1). Erregeyers (2009) suggested a normalisation of 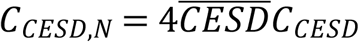, which is applied here.

We also calculate the generalised concentration index which is simply the concentration index multiplied by the mean value of CES-D, 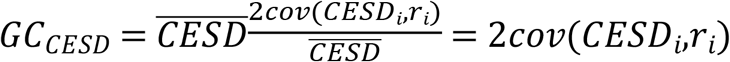.

Finally, mobility matrices are calculated for both study groups and in the case of the 1998 cohort the analysis is stratified by gender. These matrices are calculated both for the depression rate and also for the more granular CES-D8 scores.

## Results

The summary statistics presented in table 1 show the background of both study groups in terms of education (education of principal carer in the case of the 1998 cohort) and how mental health changed pre and post Covid. The 2008 cohort showed higher education generally (reflecting the fact that the principal carers are a younger cohort), and also had no “missing” responses. For both genders and both of the study groups we see a deterioration in mental health post Covid. The 1998 cohort shows nearly a doubling of the fraction with a CES-D8 score in excess of the depression threshold and also shows a higher base rate for females, with a majority of females indicating depression. The fraction of principal carers from the 2008 cohort reporting depression more than doubled, but from a considerably lower base. This data is also summarised in figures 1-2

**Table 1:** Sample characteristics and CES-D8 scores.

|  | 1998 Cohort |  |  |  | Principal Carers<br>2008 Cohort |  |
| --- | --- | --- | --- | --- | --- | --- |
|  | Females |  | Males |  |  |  |
|  | Pre<br>Covid | Post<br>Covid | Pre<br>Covid | Post<br>Covid | Pre<br>Covid | Post<br>Covid |
| **Lower**<br>**Secondary** | 0.222 |  | 0.204 |  | 0.111 |  |
| **Complete**<br>**Secondary** | 0.390 |  | 0.370 |  | 0.355 |  |
| **Diploma/Cert** | 0.157 |  | 0.178 |  | 0.221 |  |
| **Third Level** | 0.166 |  | 0.182 |  | 0.313 |  |
| **Education**<br>**missing** | 0.064 |  | 0.067 |  | 0.000 |  |
| **CES-D8**<br>**(mean)** | 4.984 | 7.561 | 3.755 | 5.828 | 2.321 | 3.978 |
| **Proportion**<br>**Depressed** | 0.307 | 0.558 | 0.225 | 0.403 | 0.107 | 0.240 |

**Figure 1:**
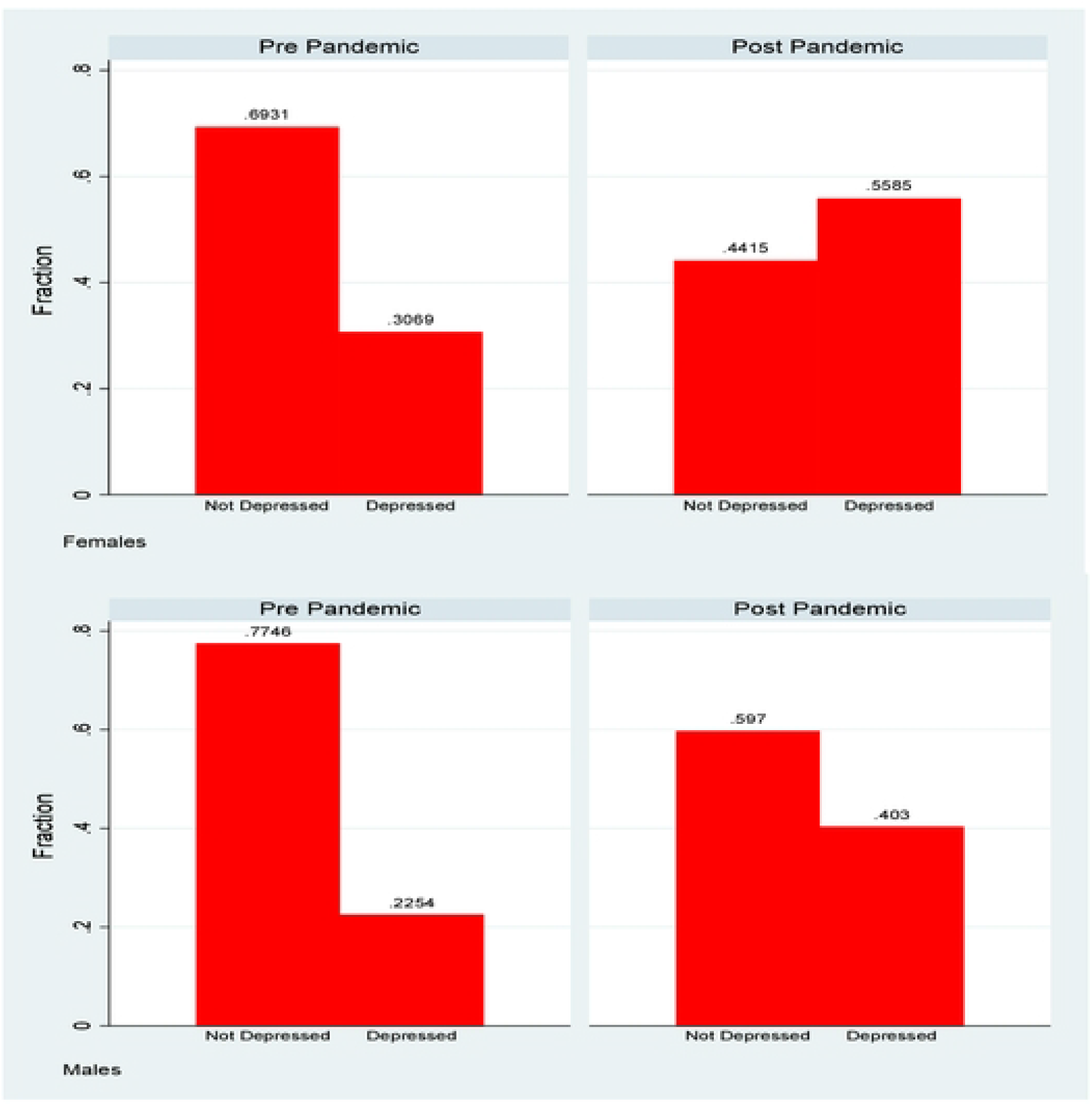
Depression rates, 1998 cohort, by gender.

**Figure 2:**
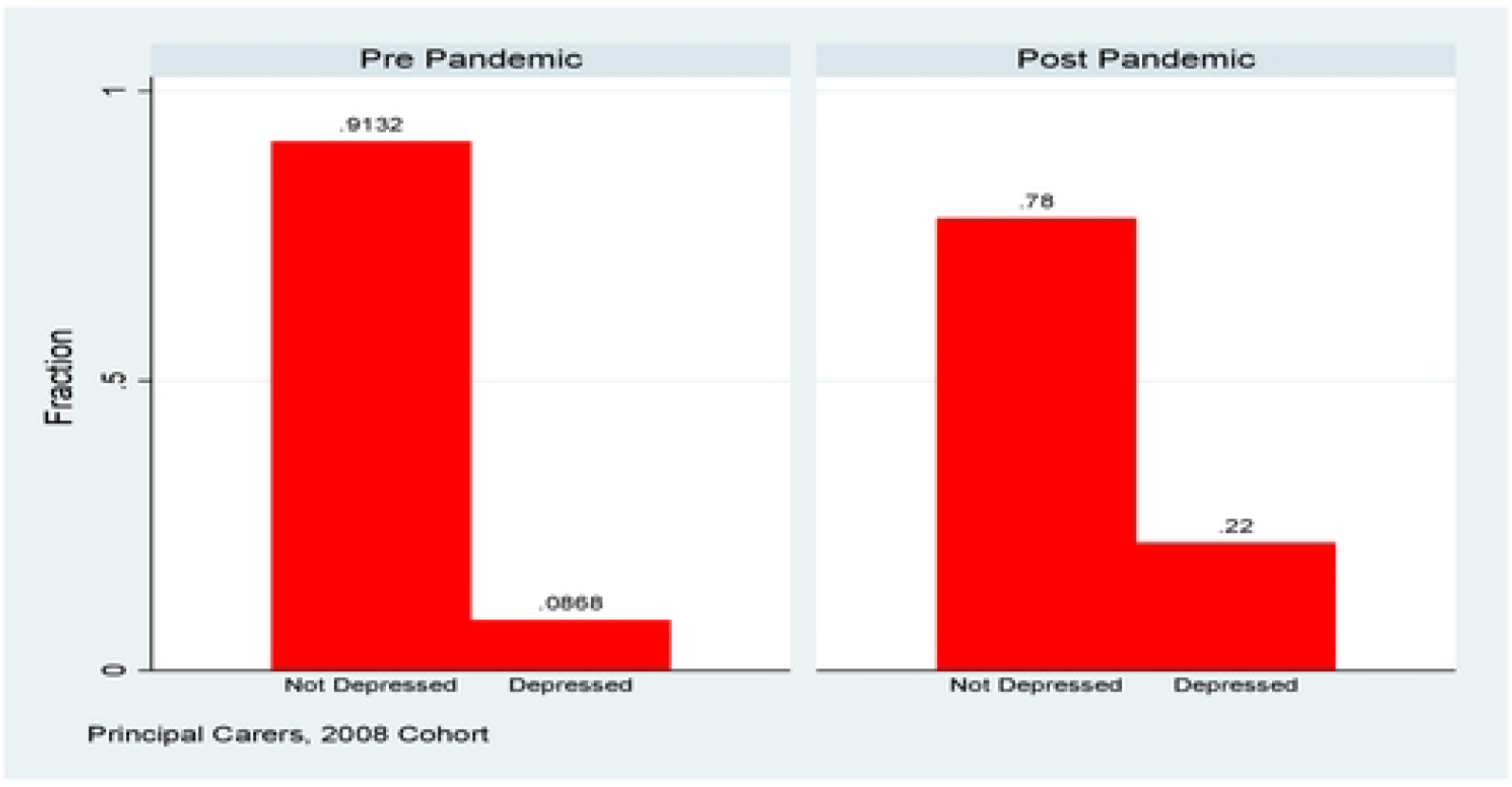
Depression rates, 2008 cohort, Principal Carers.

### Shapley Decomposition

For the 1998 cohort, the bulk of the change in the rate of depression is accounted for by a general growth in the level of CES-D8 and this is particularly so for males. For females, a change in the distribution accounts for about 20% of the increase in depression, but overall the increase in depression over the period is mostly accounted for by a general rise in CES-D8 rather than by any changes in the shape of the distribution. Note that owing to the top-censoring of the distribution at 13, the calculated mean CES-D8 is underestimated both pre and post pandemic. This will only affect the breakdown of the change in the fraction depressed if we believe that the average CES-D8 for those above 13 is *higher* post rather than pre pandemic. This seems plausible, in which case the percentage of the change in the rate of depression accounted for by overall growth as opposed to redistribution is a *lower* bound.

For the principal carers of the 2008 cohort the growth in the *level* of CES-D8 over-explains the total change. Or to phrase it another way, if we had only witnessed the change in the distribution of CES-D8 pre and post pandemic, while holding the mean constant, then depression would have *fallen*. Thus the rise in depression was completely caused by an overall deterioration in CES-D8 scores for everyone, rather than a spread in the distribution which pushed some people over the threshold.

### Socioeconomic Gradient by Maternal Education

For the 1998 cohort, the depression rates by maternal education for females show little variation, and if anything a higher rate for those with higher maternal education post covid. The figures for males are more mixed. Pre-covid, the lowest depression rates were observed for those with highest maternal education, but the relationship was not monotonic. Those with the lowest maternal education did not show the highest rates of depression. In the post-covid situation there is very little discernible gradient and depression rates for those with highest maternal education have caught up with the average for males.

For the PCs of the 2008 cohort depression rates are highest for those with the lowest education level, followed by those who have completed secondary school education. The rates for those with a diploma or cert or third level education are very close. All groups show a statistically significant increase post pandemic and the absolute differences are highest for those with the two lower levels of education. Relative rates of increase however are greater for the higher levels of education, so in relative terms at least, there is some sign of convergence in rates of depression, albeit at a higher level.

### Socioeconomic Gradient by Income

For the 1998 cohort, while the concentration index is negative for females and positive for males, in all cases it is insignificantly different from zero, indicating no socioeconomic gradient to either CES-D8 or to depression, in total, or by gender.

For the 2008 cohort of PC, the concentration index shows a fall post pandemic. However, the *generalised* concentration index i.e. the concentration index multiplied by the average level of CES-D8 shows an increase. The concentration index for the rate of depression also rises. Thus in terms of how the socioeconomic gradient has changed, the picture is quite complex. In purely relative terms the gradient is less steep post-pandemic. However allowing for the change in the average level of CES-D8 the gradient is steeper reflecting a greater concentration of higher CES-D8 values amongst the poorer and less well-educated, and this is also confirmed by the results in table 3.

**Table 2:** Shapley Decomposition for Change in Depression, Pre and Post Pandemic.

| Measure | Fraction Depression, CES-D $\geq 7$ | | | | |
| --- | --- | --- | --- | --- | --- |
|  | Pre | Post | Change | Growth Contribution (%) | Distribution Contribution (%) |
| **Total (1998)** | 0.266 | 0.481 | 0.215 | 0.172<br>80% | 0.043<br>20% |
| **Female (1998)** | 0.307 | 0.559 | 0.252 | 0.18<br>71.6% | 0.07<br>21.4% |
| **Male (1998)** | 0.225 | 0.403 | 0.178 | 0.164<br>92.1% | 0.014<br>7.9% |
| **Principal Carers (2008)** | 0.107 | 0.240 | 0.133 | 0.157<br>117.8% | -0.024<br>-17.8% |

**Table 3:** Change in Rate of Depression by Education, standard errors in brackets, *** p<0.001, **p<.01.

| **1998 Cohort** | **Depression Rate**<br>**Pre Covid** | **Depression Rate**<br>**Post Covid** | **Difference** |
| --- | --- | --- | --- |
| **Females** |  |  |  |
| *Lower Secondary* | 0.309 | 0.524 | 0.216***<br>(0.055) |
| *Complete Secondary* | 0.307 | 0.576 | 0.269***<br>(0.039) |
| *Diploma/Cert* | 0.316 | 0.574 | 0.258***<br>(0.047) |
| *Third Level* | 0.305 | 0.558 | 0.253***<br>(0.041) |
| *Missing* | 0.280 | 0.535 | 0.255**<br>(0.103) |
| **Males** |  |  |  |
| *Lower Secondary* | 0.219 | 0.394 | 0.175**<br>(0.081) |
| *Complete Secondary* | 0.261 | 0.417 | 0.156***<br>(0.034) |
| *Diploma/Cert* | 0.236 | 0.354 | 0.118**<br>(0.055) |
| *Third Level* | 0.167 | 0.426 | 0.259***<br>(0.045) |
| *Missing* | 0.178 | 0.422 | 0.245**<br>(0.104) |
| **TOTAL** | **0.266** | **0.481** | **0.215\*\*\***<br>**(0.072)** |
| **2008 Cohort**<br>**(Principal Carers)** | **Depression Rate**<br>**Pre Covid** | **Depression Rate**<br>**Post Covid** | **Difference** |
| *Lower Secondary* | 0.2188 | 0.3618 | 0.1430***<br>(0.0509) |
| *Complete Secondary* | 0.1144 | 0.2705 | 0.1561***<br>(0.0202) |
| *Diploma/Cert* | 0.0852 | 0.1973 | 0.1120***<br>(0.0182) |
| *Third Level* | 0.0747 | 0.1923 | 0.1176***<br>(0.0125) |
| **TOTAL** | **0.1071** | **0.2399** | **0.1329\*\*\***<br>**(0.0107)** |

**Table 4:** Concentration and Generalsied Concentration Indices for CES-D8 and Depression, standard errors in brackets, *** p<0.001, **p<.01.

|  | **CES-D, Concentration Index** |  | **CES-D, Generalised Concentration Index** |  | **Depression, Concentration Index** |  |
| --- | --- | --- | --- | --- | --- | --- |
|  | **Pre** | **Post** | **Pre** | **Post** | **Pre** | **Post** |
| **2008 Cohort** |  |  |  |  |  |  |
| *Females* | -0.026<br>(0.022) | -0.018<br>(0.015) | -0.128<br>(0.109) | -0.139<br>(0.112) | -0.037<br>(0.045) | -0.049<br>(0.049) |
| *Males* | 0.038<br>(0.029) | 0.009<br>(0.022) | 0.143<br>(0.110) | 0.053<br>(0.128) | 0.020<br>(0.048) | 0.055<br>(0.057) |
| **Total** | -0.003<br>(0.013) | -0.011<br>(0.013) | -0.015<br>(0.080) | -0.072<br>(0.087) | -0.015<br>(0.033) | -0.007<br>(0.038) |
| **1998 Cohort** |  |  |  |  |  |  |
| **Total** | -0.107***<br>(0.021) | -0.069***<br>(0.014) | -0.248***<br>(0.049) | -0.275***<br>(0.053) | -0.106***<br>(0.020) | -0.137***<br>(0.025) |

### Transitions in mental health pre and post Covid

As well as looking at overall rates of depression, we also look at mobility over the course of the pandemic. The *tabplot* command in Stata offers a visually intuitive way of capturing this. It combines a transition matrix with a histogram, so we can easily see the transitions into and out of depression. Below we present this for the 22/23 year olds from the 1998 cohort, where depression is defined as a CES-D score greater than or equal to 7.

Figure 6 shows that of the 73.3 per cent of the sample (45.7 per cent + 27.6 per cent) of those who were not depressed pre-Covid, 27.6 percent transitioned into depression post-Covid. Of the 26.7 per cent who were depressed pre-Covid, 6.2 per cent transitioned back into non-depression but 20.5 per cent remained depressed.

Figure 7 shows higher percentages of females depressed pre Covid and then also a higher rate of transition into depression post Covid. Over 31 per cent of females become depressed post Covid as opposed to 24 per cent of males.

Figure 8 the equivalent graphs for PCs from the 2008 cohort. What is perhaps most notable here is the relatively low rate of transition out of depression over the pandemic, in contrast to the 1998 cohort, perhaps reflecting lesser volatility of mental health.

It is also possible to examine mobility using these transition matrices at a more granular level. The matrix below shows transitions across different values of CES-D8 for the 1998 cohort. In examining these graphs there are a number of factors to bear in mind. Transitions which happen below the critical value of 7 are arguably of less interest since they do not involve a transition across the depression threshold. Because the data are truncated at values of 13 we do not observe transitions from say a value of 14 to 17, they are all absorbed in the category “>=13”. Finally, we cannot assign any cardinality to these transitions e.g. we cannot say that a move from 2 to 4 represents the same change as a move from 4 to 6, or from 8 to 10. Nevertheless, it seems reasonable to suggest that a move from 2 to 12 represents a greater deterioration in health than a transition from 2 to 8, or from 1 to 8.

We divide the matrix into four quadrants. The north-west quadrant represents people who are not depressed either pre or post Covid, so in some sense they are of less concern. While a move to the right within this quadrant is better avoided as it indicates a deterioration in mental health, since it does not involve a move across the threshold it is arguably of less concern than moves which *do* cross the threshold.

The south-west quadrant is the quadrant of good news as it comprises people whose mental health improved over the pandemic and who moved from depressed to non-depressed. Alas, there are relatively few of those. The north-east and south-east quadrants are of most concern, and the advantage of the detailed transition matrix is that it does not just tell us that people crossed into depression (or stayed depressed) but it also gives some idea (allowing for the caveat re cardinalisation mentioned above) of *how much* someone’s mental health deteriorated. Given this, it is disturbing to note that if we look at all the columns in the north-east quadrant, the most heavily populated is the right-most one, where the post-Covid CES-D8 score is highest. In other words, for those who transitioned into depression, excepting those who had a pre-Covid CES-D score of 0 or 1, for all the others a higher proportion moved to a post-Covid score of 13 or over, the highest (worst) score achievable, than any other score. Mental health did not just deteriorate, it seems to have deteriorated quite significantly.

Finally, in the south-east quadrant of people who were depressed pre and post Covid it is also noticeable that the right-most column is the most heavily populated. People whose mental health was above the depression threshold saw further deterioration, in many cases to the highest (worst) recorded CES-D8 score.

We also show the detailed transition matrices by gender in figure 10. What is probably most noticeable about these matrices is that the phenomenon of moving to the worst state is clearly more pronounced for females than for males. To use an informal metric, of the columns in the north-east quadrant for females in 6/7 cases the most populated is the right-most one, while for males it is only 3/7. There was a deterioration in mental health for males but it seems to have been much less severe than was the case for females.

Finally figure 11 shows the same detailed transition matrix for mothers of the 2008 cohort. As is already evident in the less detailed matrix looking just at depression, the overall level of depression and of transition into depression is less than is the case for the 1998 cohort. The “heavy rightmost column” phenomenon in the north-east quadrant also does not seem to be present. In no case is the right-most column of the north-east quadrant the most heavily populated. While the mental health of the PCs of the 2008 cohort did deteriorate, the deterioration seems to have been less severe than in the case of the 22/23 year olds of the 1998 cohort.

## Discussion

This study examines the evolution of mental health in Ireland pre and post the Covid 19 pandemic for two distinct groups. First of all a cohort of young adults born in 1997-98 and secondly a group of PCs (predominantly female) of a cohort of children born in October 2008. The particular contribution of this study is the availability of a consistent measure of mental health (the CES-D8) before and after the pandemic for the same group of people, thus controlling for time invariant characteristics.

The study finds that mental health deteriorated for both these groups. The deterioration was most pronounced among females of the 1998 cohort. Their CES-D8 scores pre-pandemic were the highest (indicating worse mental health) and they also showed the greatest deterioration post pandemic, to such an extent that the majority of females of the 1998 cohort had a score at or above the key threshold of 7, thus indicating depression. Depression rates for males from the 1998 cohort and for the principal carers of the 2008 cohort also showed increases, though the level of depression for the latter group was considerably lower, at around 22 per cent.

Our analysis also sheds light on how the rate of depression (as measured by the fraction of the sample with a CES-D8 score greater than or equal to 7) evolved. Effectively we are looking at how the mass of the distribution to the right of a specific threshold changes over time. This can happen because the whole distribution shifts to the right (the “growth effect”) while retaining the same shape, or because the shape of the distribution changes (the “distribution effect”) or both. Our results show that for both males and females from the 1998 cohort that the growth effect dominated, accounting for between 70 and 90 pe cent of the overall increase. Thus the increase in the rate of depression was mostly caused by an overall decline in mental health, rather than a decline for those near the threshold. The dominance of the growth effect is even greater for the principal carers of the 2008 cohort, where it accounts for more than 100 per cent of the change.

Our analysis also examines how the distribution of mental health with respect to measures of socioeconomic status (SES) evolved pre and post pandemic. For many health conditions a “gradient” is observed where those with lower SES (whether measured by income, social class or education) have poorer health outcomes (Kakwani et al, 1997).We use household after tax income and maternal education as measures of SES for the 1998 cohort and actual educational attainment and after tax household as measures for the PCs of the 2008 cohort. Figures 3-4 show how depression rates change by maternal education for the 1998 cohort and figure 5 shows this information by education for the PCs of the 2008 cohort, with the underlying numbers in table 3.

**Figure 3:**
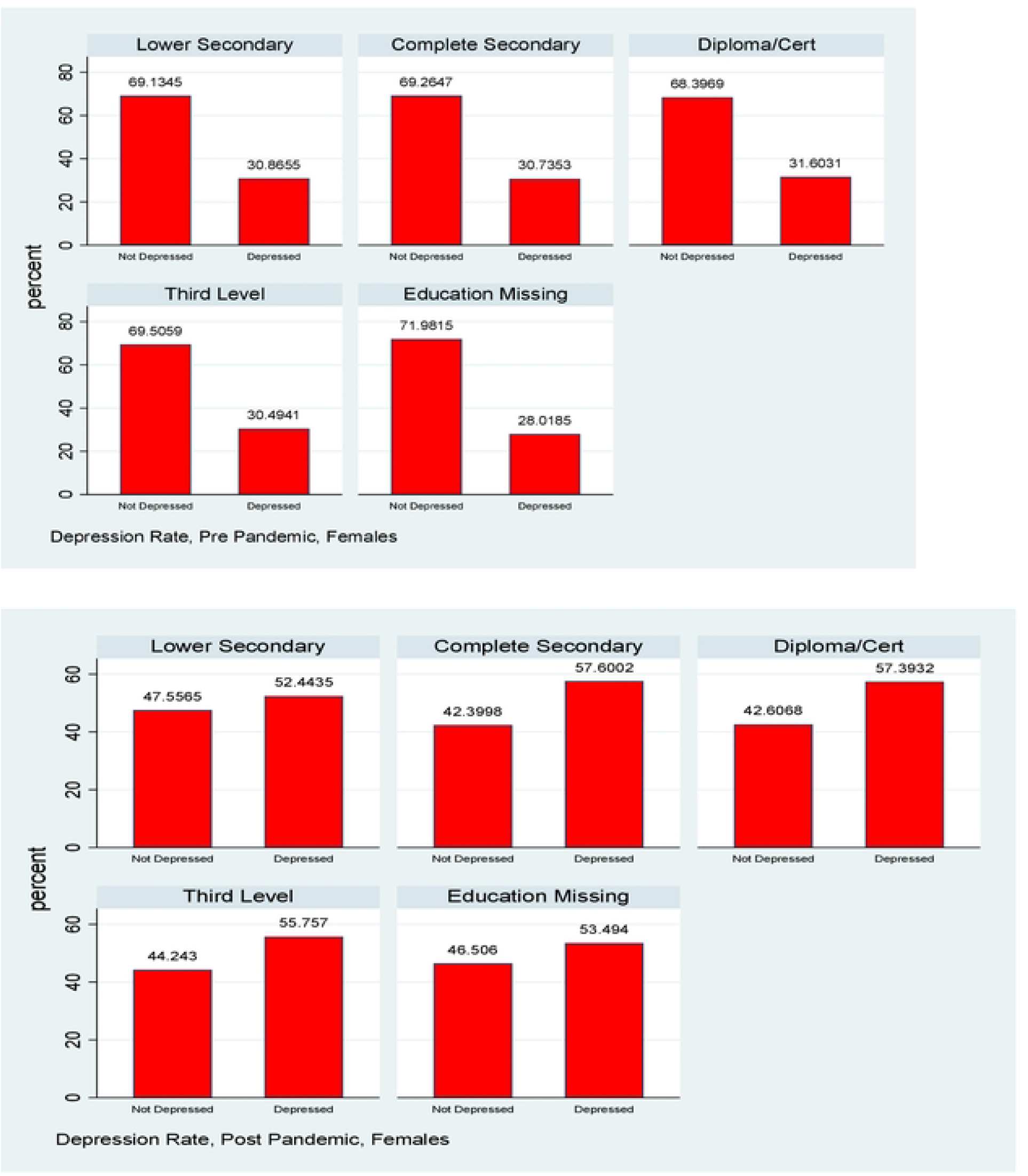
Depression Rates Pre and Post Pandemic, 1998 Cohort, Females, by maternal education.

**Figure 4:**
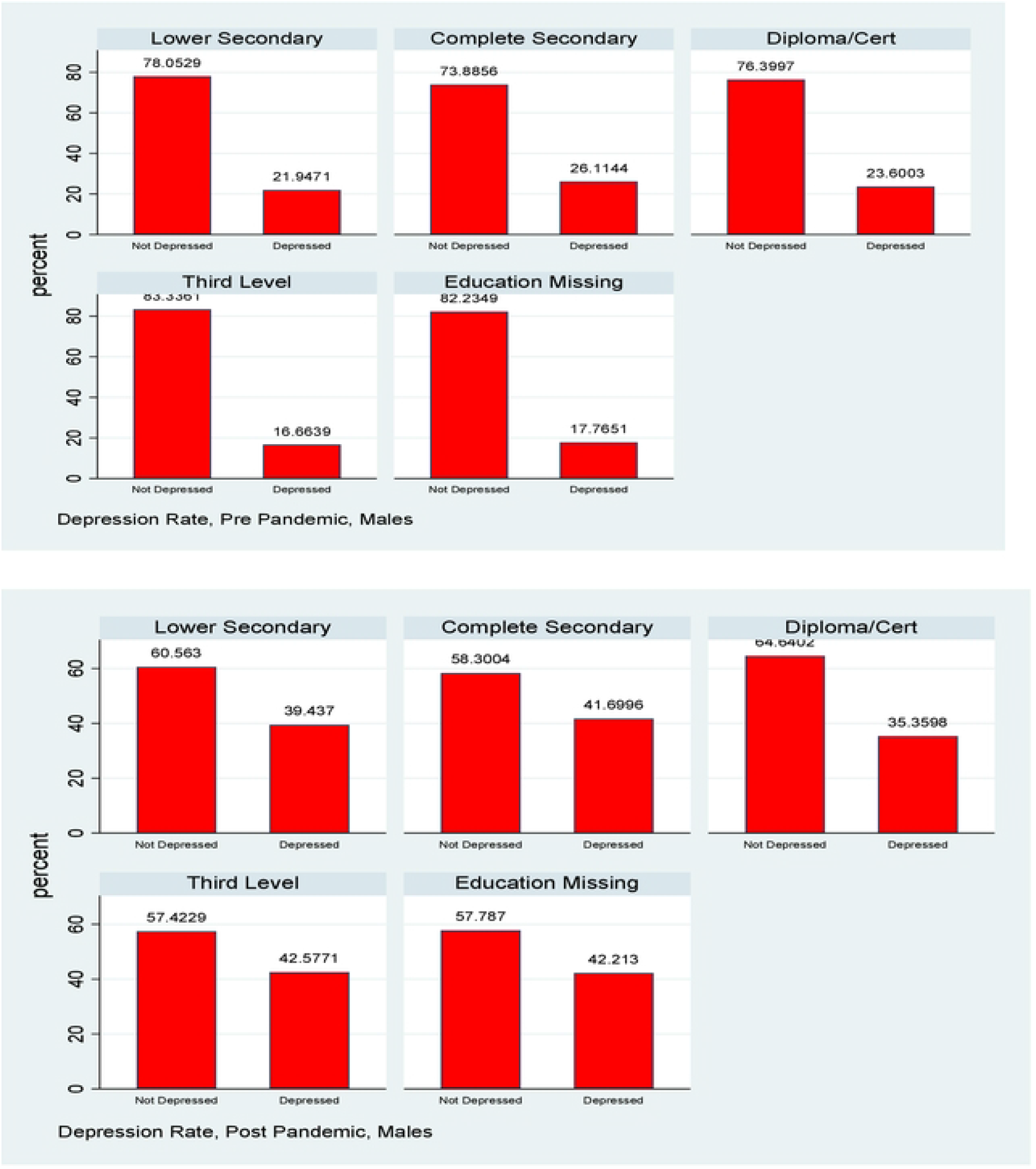
Depression Rates Pre and Post Pandemic, 1998 Cohort, Males, by maternal education.

**Figure 5:**
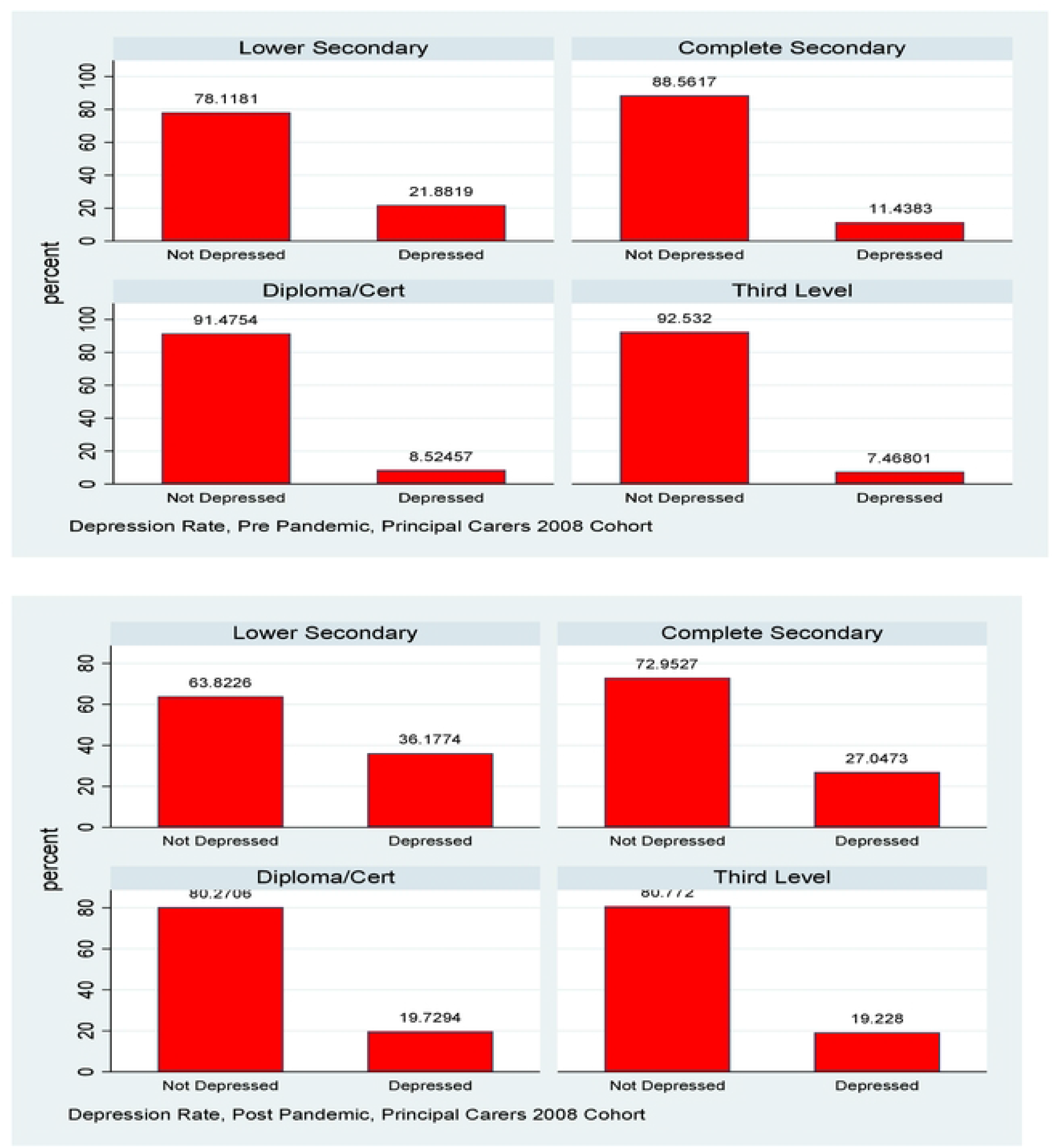
Depression Rates Pre and Post Pandemic, 2008 Cohort, Principal Carers, by education.

For females in the 1998 cohort there is little evidence of a gradient in depression by maternal education either pre or post pandemic, though of course the overall rate of depression has increased. For males there is some evidence of a slight gradient pre pandemic with depression rates lower for those whose mothers have post secondary school education, but this has mostly vanished post pandemic. Again, the overall rate of depression increased but it is noticeable that there is very little evidence of a gradient by SES, and, whatever slight one did exist, disappeared with the pandemic.

Figure 5 shows a discernible gradient in depression rates by education both pre and post pandemic for the PCs of the 2008 cohort. The relative gaps by education compress slightly post pandemic, but since absolute levels are considerably higher the absolute gaps have widened.

Results are also presented for these gradients by income and in this case, given that income is a continuous cardinal variable it is possible to calculate a summary measure, the concentration index. Clearly income and education are not equivalent but they do tend to be quite highly correlated so it is no surprise to see that the results by education are broadly replicated. For the 1998 cohort there is no statistically significant gradient for either gender, pre or post pandemic. For the PCs of the 2008 cohort, when measured on a purely relative basis we see a slight fall in the concentration index, but when allowing for the increase in the overall rate of depression, the index increases slightly. Overall, however, it seems fairest to say that the pandemic had little effect on what SES gradients in mental health already existed pre pandemic.

Our final analysis specifically looks at mobility, exploiting the longitudinal nature of the data. Again, it is instructive to stratify the analysis by gender for the 1998 cohort. Reflecting the changes in the overall rates of depression, there are more entries into depression rather than exits from depression and this is more pronounced for females than males in the 1998 cohort, which in turn is more pronounced than for the PCs of the 2008 cohort. Perhaps the greatest difference is seen however when we look at the more granular transition matrices in figures 10-11. The data on CES-D8 is truncated at a value of 13, so we cannot observe transitions to the very highest values (and hence worst mental health outcomes). However CES-D8 values of 13 or greater clearly indicate more severe depression than values just above the threshold of 7. In this regard it is striking that of those females from the 1998 cohort who transition into depression, a noticeably higher fraction record values of 13 or greater than is the case for males or for the PCs from the 2008 cohort. Thus it seems fair to say that for the three distinct groups we observe in this study, not only did females from the 1998 cohort see a greater increase in the rate of depression over the pandemic, but they also seem to have moved to a more intense level of depression.

**Figure 6:**
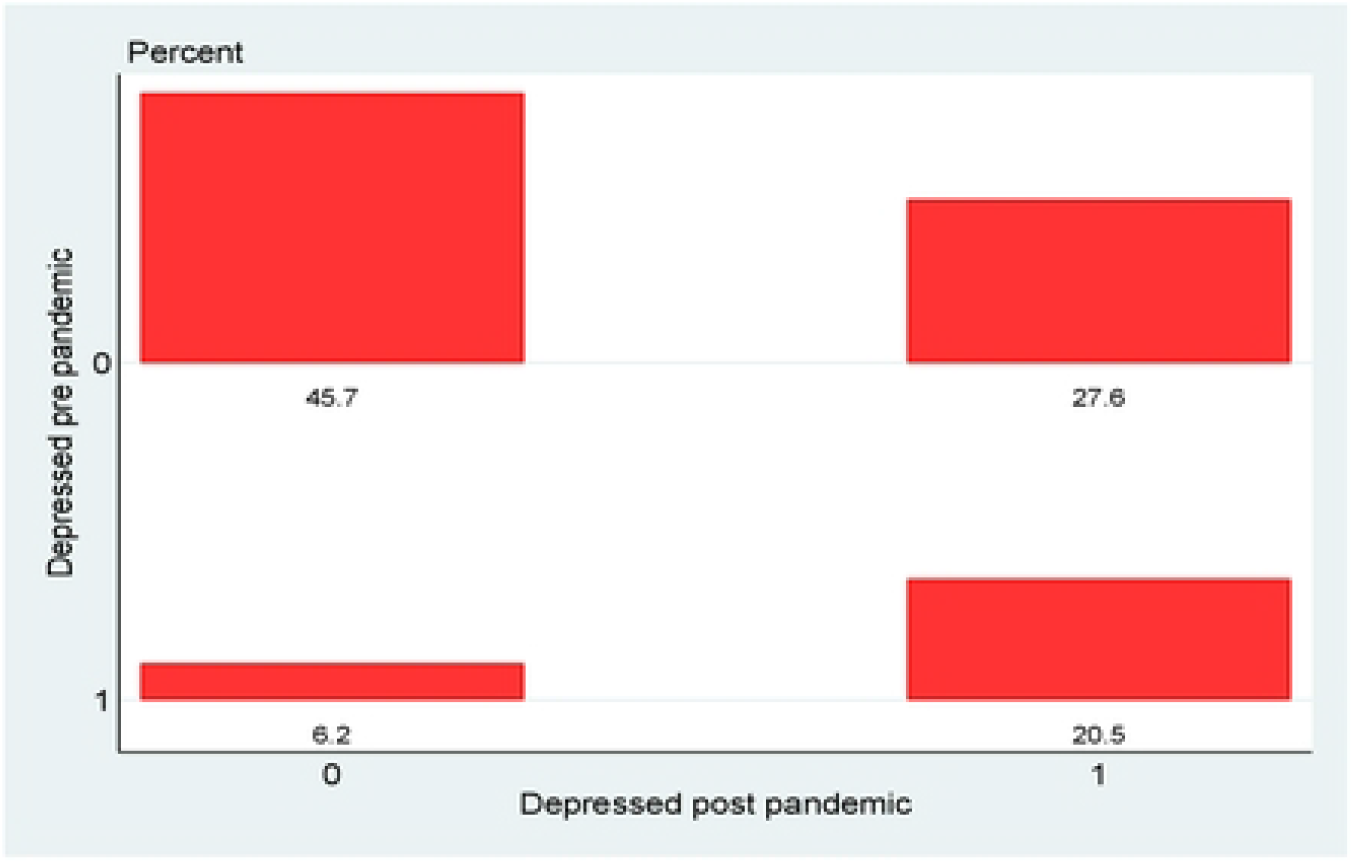
Transition matrix, 1998 cohort, total.

**Figure 7:**
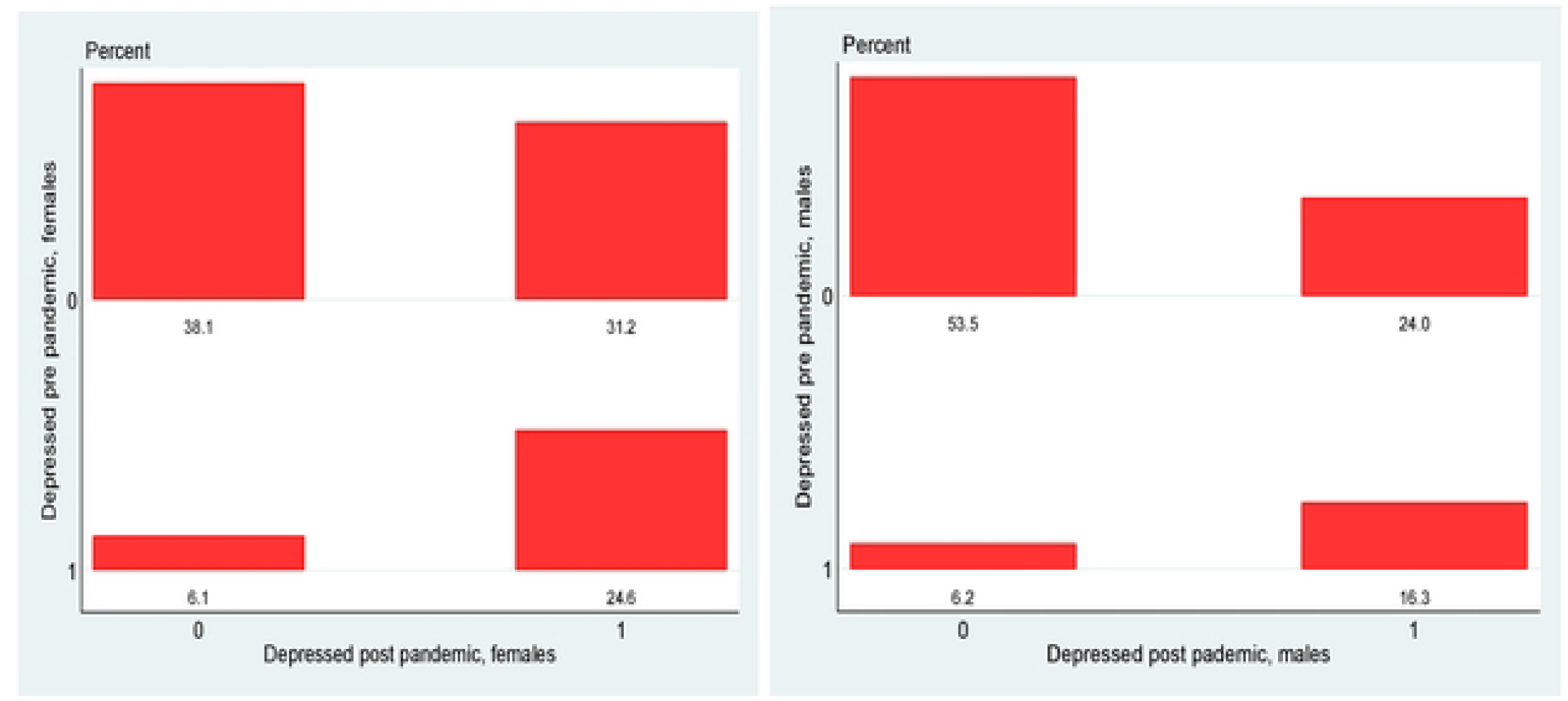
Transition matrix, 1998 cohort, by gender.

**Figure 8:**
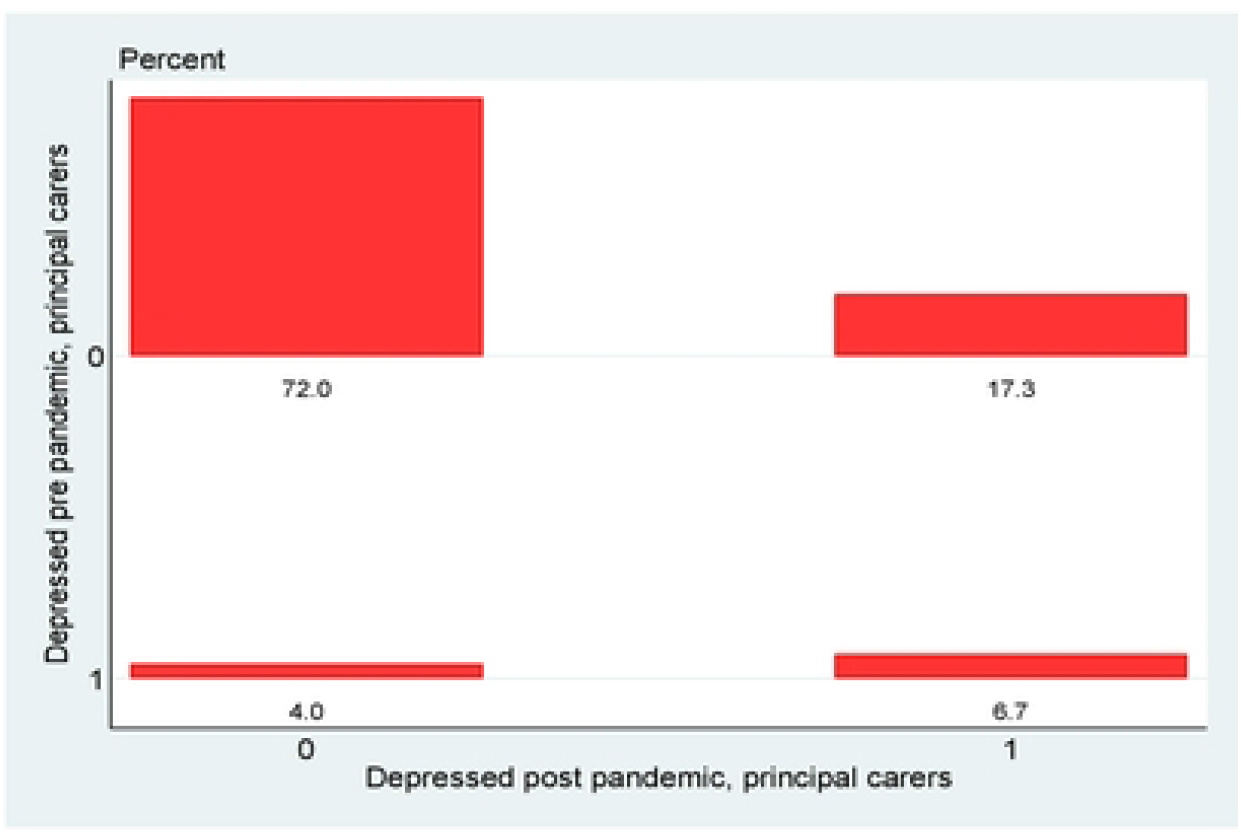
Transition matrix, 2008 cohort, principal carers.

**Figure 9:**
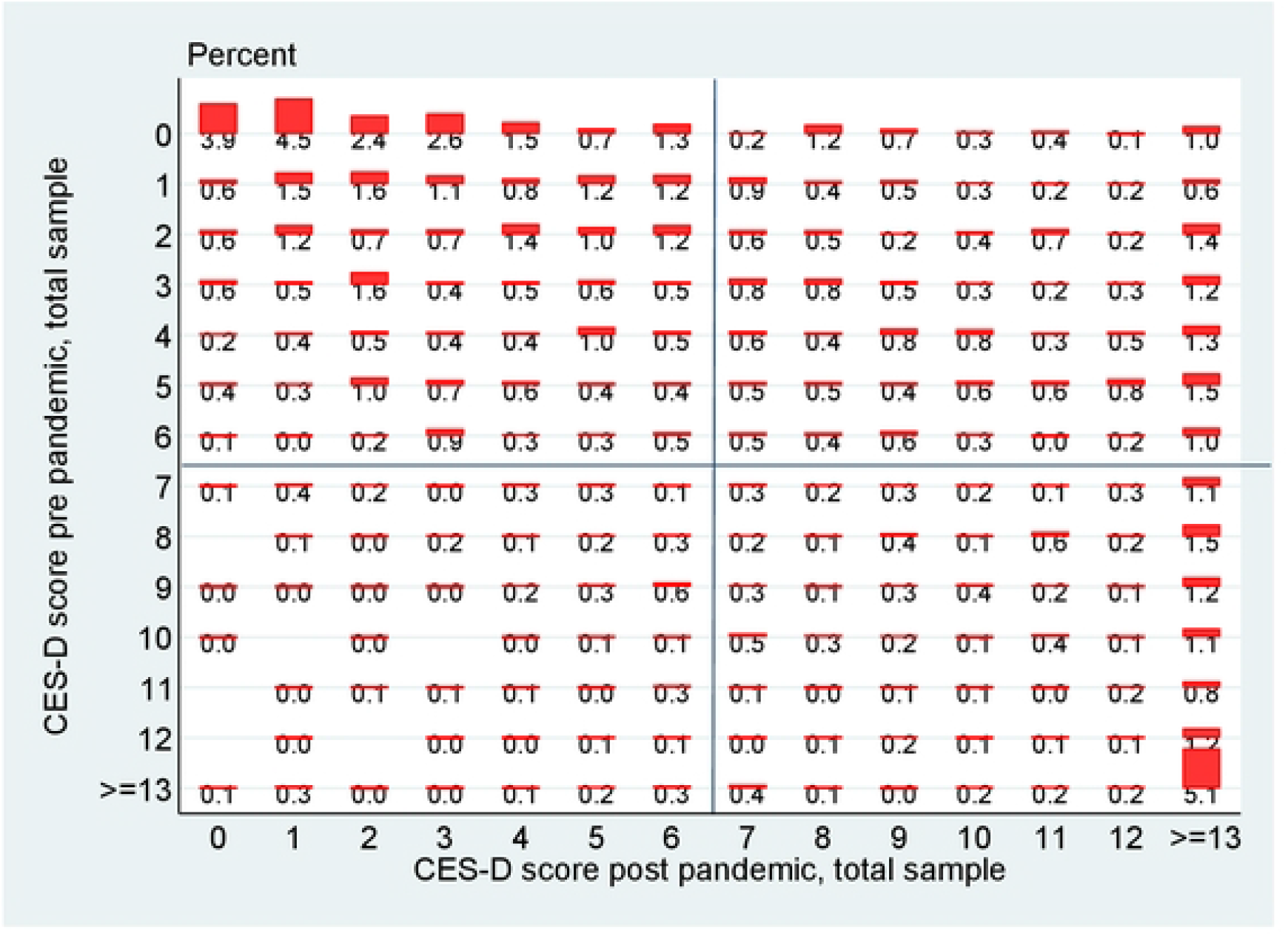
Transition matrix, 1998 cohort, total sample, detailed.

**Figure 10:**
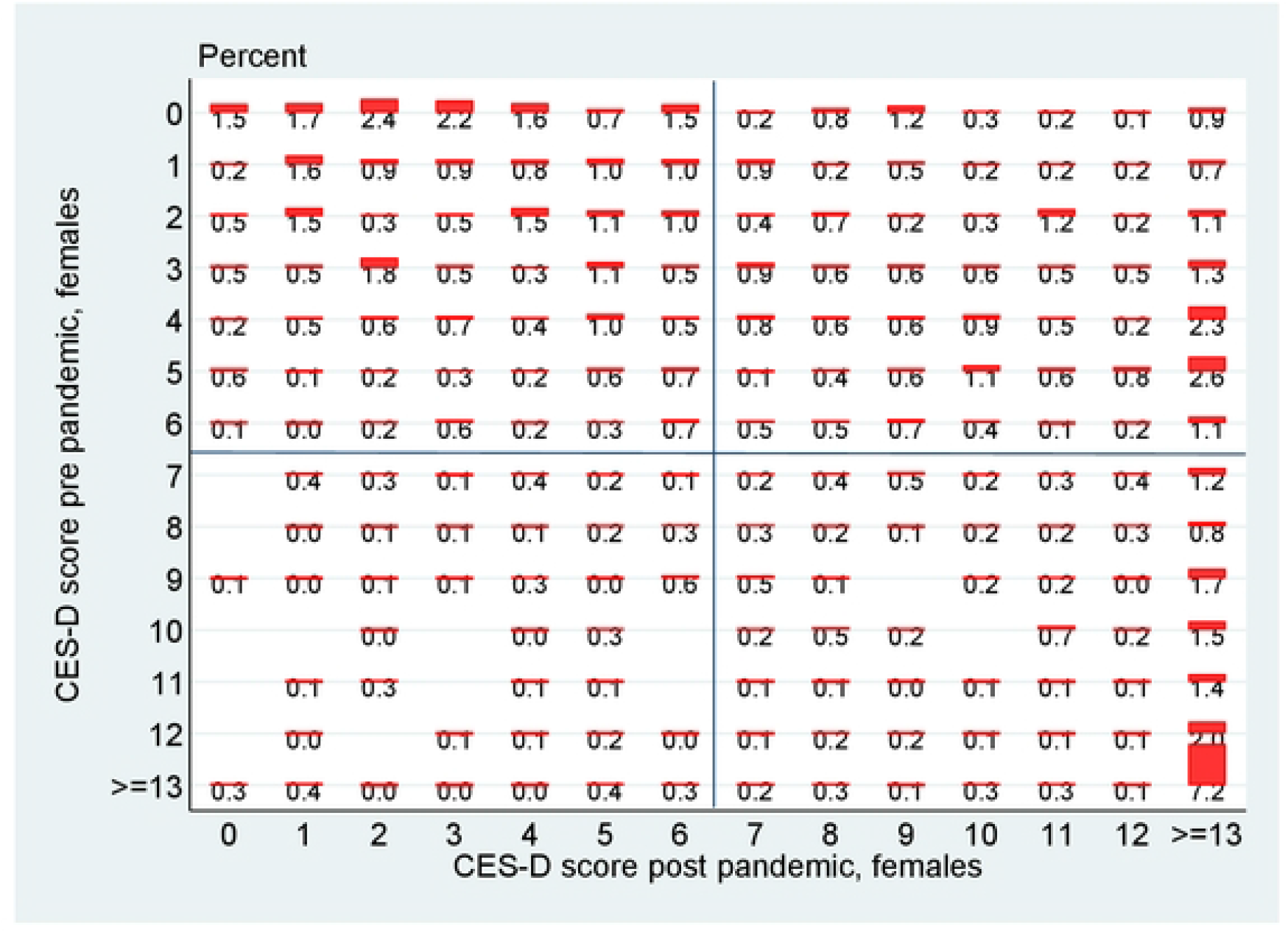
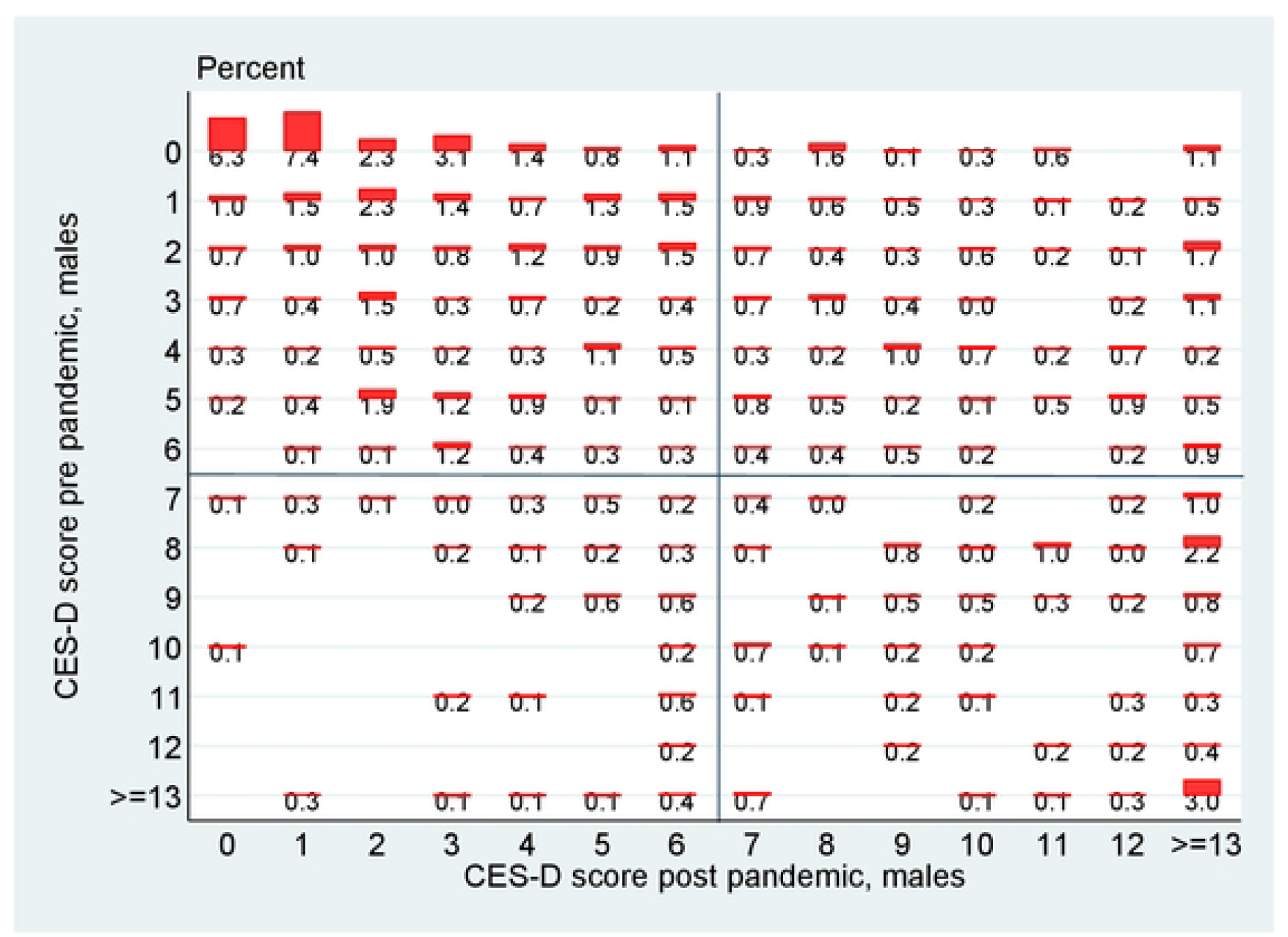
Transition matrices, 1998 cohort, detailed, by gender for females.

**Figure 11:**
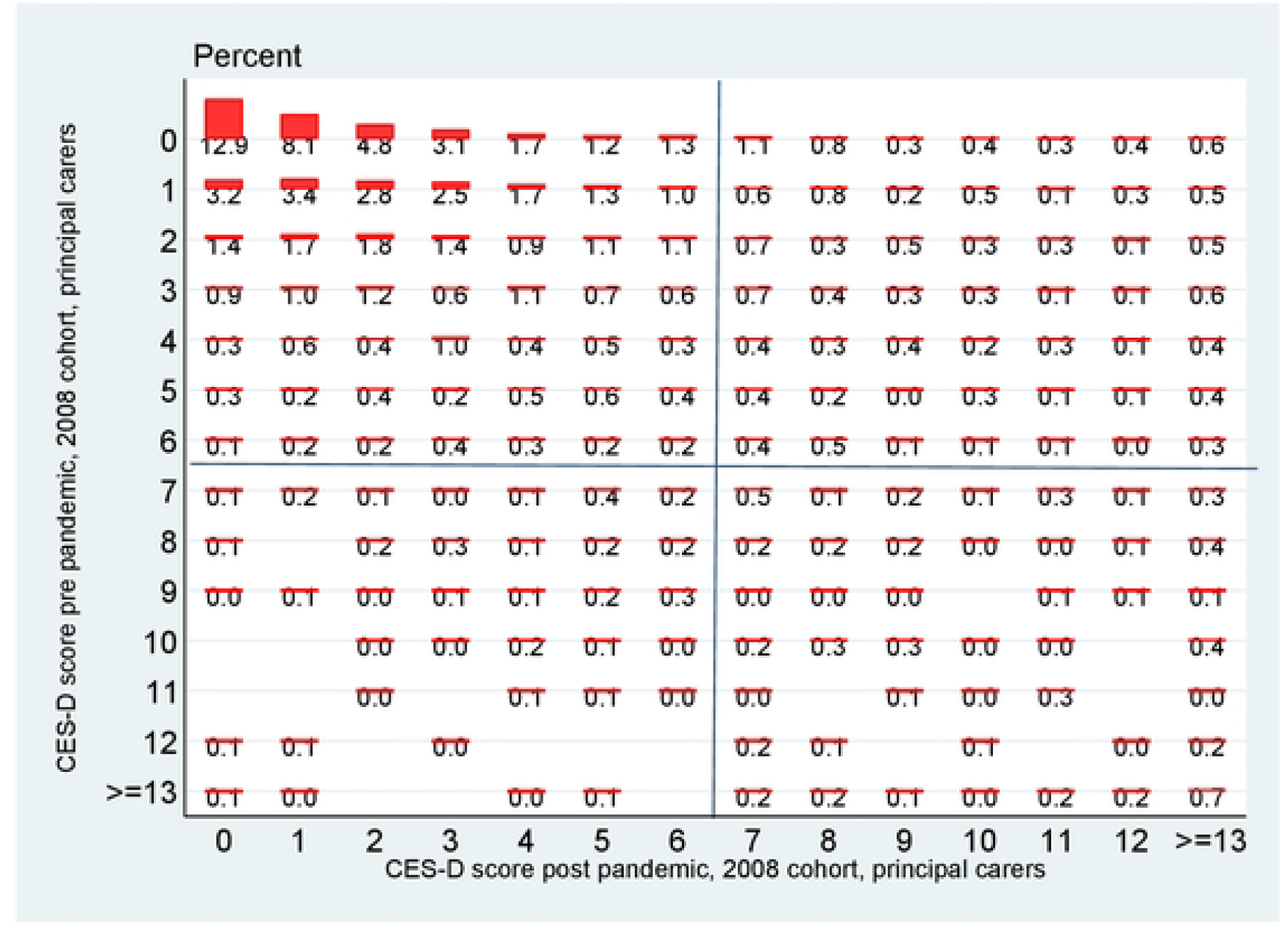
Transition matrix, 2008 cohort, principal carers, detailed.

It is important also to acknowledge some potential caveats with the analysis. Thus it is possible that the increase in depression for the 1998 cohort over the years leading up to December 2020 could be to some degree explained by age. As Banks and Xu (2020) show in their study of mental health in the UK during the early stages of the pandemic, well-being tends to show a U shaped relationship with age, with fairly steep changes in mental health problems for younger people, and also for those entering retirement. They also show that there is a seasonal aspect to mental health, and that it tends to deteriorate going into winter months. The GUI Covid surveys were held in December 2020 and evidence from the UK Household Longitudinal Study (UKHLS) suggests that seasonal factors are significant for this month. Banks and Xu use the rich data available in the UKHLS to estimate a counterfactual of how mental health might have evolved in the absence of the pandemic and their analysis for the group comparable in age to the 1998 cohort suggests that the pandemic accounted for about 75% of the observed deterioration in mental health, with age-related developments accounting for the remainder. Unfortunately GUI lacks a consistent measure of mental health over a sufficient sequence of waves to permit the construction of a similar counterfactual, but this issue should be borne in mind when interpreting our results. The lack of information on the month of interview for the pre pandemic survey also prevents us from taking account of potential seasonal factors.

In terms of the mobility analysis, since we only observe two data points (the surveys pre and post pandemic) it is possible that we do not observe transitions into and out of mental health which happen *between* these surveys. Thus a transition into (or out of) depression may have occurred after the pre pandemic survey but *before* the arrival of Covid, perhaps for the age related reasons referred to above. It is also possible that an observation may be observed to have no transition yet may have transferred for example into and back out of depression.

In conclusion, this study examines the evolution of mental health as measured in December 2020, nine months into the pandemic compared to observations pre pandemic for two cohorts of people. A deterioration in mental health was observed for both cohorts and particularly for younger women. The increase in the rate of depression predominantly occurred due to an overall decline in mental health rather than being concentrated amongst those already vulnerable (in the sense of being near the depression threshold). There was little, if any, change in the socioeconomic gradient associated with mental health and virtually no gradient at all was observed pre or post pandemic for the 1998 cohort. Finally, mobility analysis revealed that not only did females from the 1998 cohort show greater transitions into depression, they also appeared to transition into more extreme levels of depression. This may be partly due to age related changes which would have occurred anyway in the absence of the pandemic. It remains to be seen what at what level mental health will eventually stabilise post pandemic.

## Data Availability

Data are held in a public repository:The Irish Social Science Data Archive. https://www.ucd.ie/issda/data/guicovid19/

## Acknowledgement

I would like to acknowledge helpful comments from Orla Doyle and participants at an internal seminar in University College Dublin. The usual disclaimer applies.

## Notes

### Competing Interest Statement

The authors have declared no competing interest.

### Funding Statement

The author received no specific funding for this work

